# Impaired Th17 immunity in recurrent *C. difficile* infection is ameliorated by fecal microbial transplantation

**DOI:** 10.1101/2020.06.05.20114876

**Authors:** Laura Cook, William D. Rees, May Q. Wong, Xiaojiao Wang, Hannah Peters, Laura Oliveira, Torey Lau, Regan Mah, Brian Bressler, Rebecca Gomez, I-Ting Chow, Eddie A. James, William W. Kwok, Megan K. Levings, Theodore S. Steiner

## Abstract

**Background & Aims:** *Clostridioides difficile* is a leading cause of infectious diarrhea and an urgent antimicrobial resistant threat. Symptoms are caused by its toxins, TcdA and TcdB, with many patients developing recurrent *C. difficile* infection (CDI), requiring fecal microbiota transplant (FMT). Antibody levels have not been useful in predicting patient outcomes, which is an unmet need. We aimed to characterize T cell-mediated immunity to *C. difficile* toxins and assess how these responses were affected by FMT.

**Methods:** We obtained blood samples from patients with newly acquired CDI, recurrent CDI (with a subset receiving FMT), inflammatory bowel disease with no history of CDI, and healthy individuals (controls). Toxin-specific CD4^+^ T cell responses were analysed using a whole blood flow cytometry antigen-induced marker assay. Serum antibodies were measured by ELISA. Tetramer guided mapping was used to identify HLA-II-restricted TcdB epitopes and DNA was extracted from TcdB-specific CD4^+^ T cells for TCR repertoire analysis by Sanger sequencing.

**Results:** CD4^+^ T cell responses to *C. difficile* toxins were functionally diverse. Compared to controls, individuals with CDI, or inflammatory bowel disease had significantly higher frequencies of TcdB-specific CD4^+^ T cells. Subjects with recurrent CDI had reduced proportions of TcdB-specific CD4^+^ Th17 cells, FMT reversed this deficit and increased toxin-specific antibody production.

**Conclusions:** These data suggest that effective T cell immunity to *C. difficile* requires the development of Th17 cells. In addition, they show that an unknown aspect of the therapeutic effect of FMT may be enhanced T and B cell-mediated immunity to TcdB.

GRAPHICAL ABSTRACT

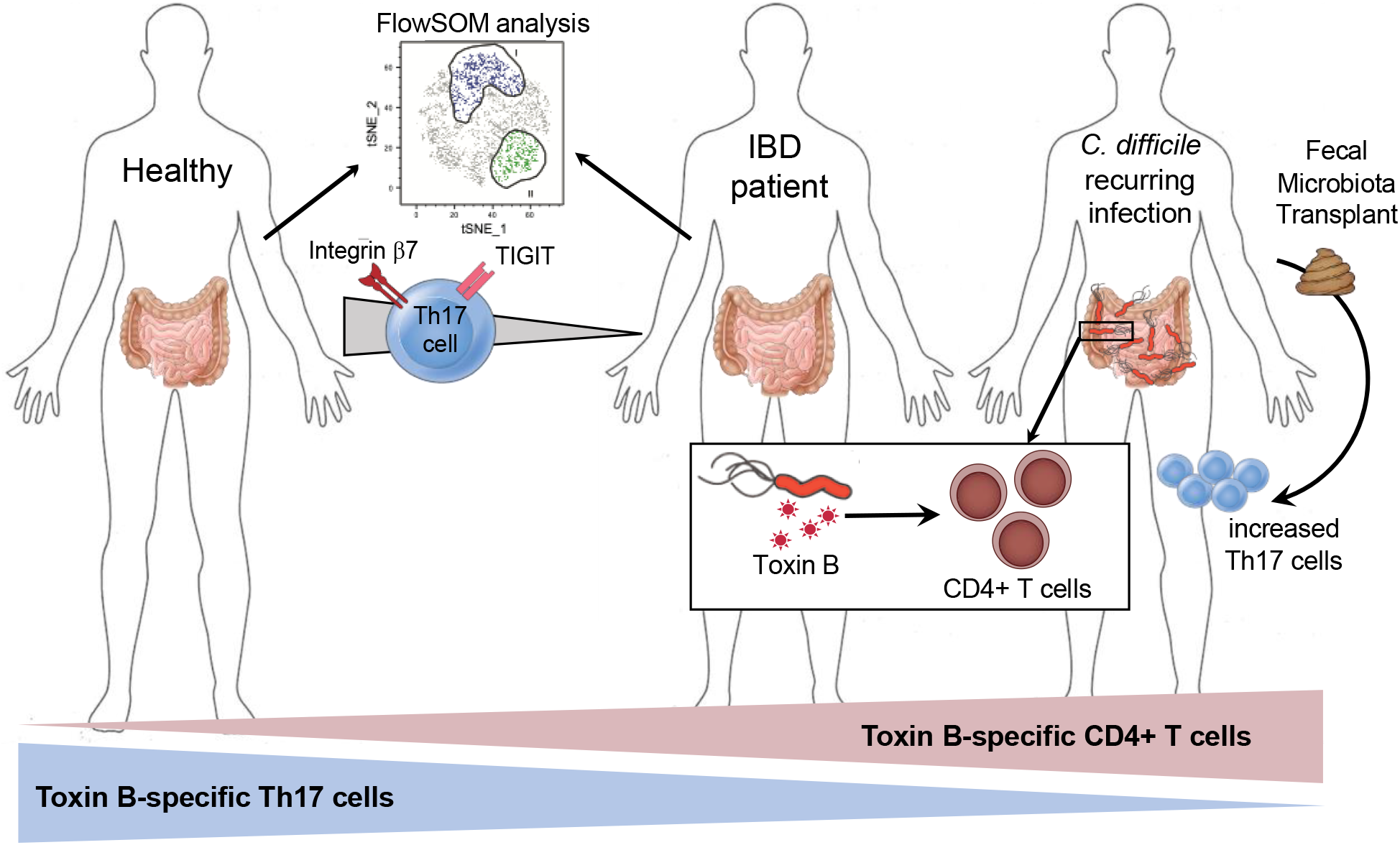

## INTRODUCTION

*Clostridioides difficile* is a gram positive, toxin-producing bacteria that is a major cause of hospital-acquired infection ^1^. *C. difficile* infection (CDI) is the leading cause of gastroenteritis-associated death in North America ^2^. Although most people are colonized with *C. difficile*, its growth is normally kept in check by commensal bacteria. However, when commensal bacteria are altered, for instance following antibiotic treatment, *C. difficile* can replicate and produce its two pathogenic toxins: TcdA and TcdB ^3^. These toxins cause intestinal epithelial injury and inflammation, resulting in CDI symptoms including diarrhoea and abdominal pain ^3^.

First line CDI treatment is typically metronidazole or vancomycin, antibiotics which kill vegetative forms and eliminate toxin production ^4^. However, in 25-35% of patients commensals fail to effectively repopulate the colon, allowing antibiotic-resistant *C. difficile* spores to germinate and cause recurrent disease ^5^. Fecal microbiota transplant (FMT) is an effective second-line therapy for recurrent CDI ^6^, but it remains difficult to access, has infectious risk ^7^, and multiple treatments may be needed for success. Newer treatments include the humanized monoclonal IgG bezlotoxumab (anti-TcdB), which partially protected against recurrent infection in two phase III trials ^8^, although due to cost it is desirable to prioritise the treatment of high-risk patients. At present there are no reliable clinical tools to predict disease recurrence.

Individuals with inflammatory bowel disease (IBD) have increased risk of CDI ^9^, which prevents mucosal healing and exacerbates long-term complications of IBD. The role of *C. difficile* in the ongoing gut dysbiosis and inflammation in IBD patients remains undetermined and likely underappreciated due to overlapping symptoms and the lack of testing that can reliably distinguish *C. difficile* colonization from active CDI ^10^.

Despite the prevalence of CDI, the immune response to this pathogen is poorly characterized, with anti-toxin antibody levels having limited clinical use ^11,12^. Therefore, we investigated the human CD4^+^ T cell response towards TcdA and TcdB following natural infection with *C. difficile*. We aimed to identify distinguishing features of CD4^+^ T cell immunity in patients with primary versus recurrent CDI, as well as in patients with inflammatory bowel disease but no clinical history of CDI. We also investigated how FMT affects CDI immunity to gain a better mechanistic understating of why this is an effective therapy in recurrent CDI.

## MATERIALS AND METHODS

### Subjects

Study protocols were approved by Clinical Research Ethics Boards of the University of British Columbia (H15-01682, H09-01238 and H18-02553) and the Vancouver Coastal Health Authority (V15-01682). All participants were ≥18 years old and provided written informed consent. Inclusion criteria for CDI cohorts was a laboratory-confirmed *C. difficile* infection (CDI) diagnosed by a positive PCR assay for *tcdB* in the presence of diarrhea (≥ 3 loose or watery bowel movements per day for ≥ 2 consecutive days). Exclusion criteria for all cohorts were HIV infection, receipt of IVIG or rituximab within 6 months, hemorrhagic disorder precluding phlebotomy, or pregnancy. Details of cohorts are in **Table 1**.

**Table 1.** Patient characteristics of recurring and new onset CDI and healthy controls.

| **Cohort** | **n** | **Age (years)** | **Sex** | **Treatment** | **Outcome** |
| --- | --- | --- | --- | --- | --- |
| New Onset CDI | 31 | Median: 53<br>Range: 20-95 | 12/31 (39%)<br>Female | Vancomycin (8)<br>Metronidazole (22)<br>Other (1) | Not available, cohort still being followed. |
| Recurrent CDI | 39 | Median: 57<br>Range: 18-95 | 28/39 (72%)<br>Female | Vancomycin (29)<br>Metronidazole (4)<br>Other (5)<br>Untreated (1) | Self-cure (2)<br>Cured with antibiotics (7)<br>Cured with FMT (21)<br>Not cured or died (6)<br>Unknown (3) |
| FMT analysis cohort | 22 | Median: 61<br>Range: 18-95 | 16/22(68%)<br>Female | Vancomycin (22) | Cured with 1-2 FMT (20)<br>Recurrent infection (1)<br>Unknown (1) |
| Healthy controls | 21 | Median: 47<br>Range: 27-84 | 11/21 (52%)<br>Female | NA | NA |

<u>New onset CDI cohort:</u> patients with a primary incidence of CDI receiving physician-prescribed, metronidazole or vancomycin, samples collected within 5 days of diagnosis.

<u>Recurrent CDI cohort:</u> patients receiving assessment and/or treatment for recurrent CDI with ≥ 1 episode of CDI and ≥ 1 unprovoked recurrence of CDI (return of diarrhea within two months after successful treatment for CDI, without additional antibiotics). Blood collected prior to FMT.

<u>FMT analysis group:</u> subset of recurrent CDI cohort, second blood draw 8-12 weeks following FMT. FMTs were administered via a single 50 mL enema from healthy donor stool, frozen and thawed as reported ^13^ under approved UBC ethics protocols (H13-01221 and H15-00763).

<u>IBD cohort:</u> established diagnosis of either Crohn’s disease (CD) or ulcerative colitis (UC) and age and sex-matched healthy controls. Clinical data have been previously published ^14^.

<u>Healthy controls:</u> no previous diagnosis of CDI, immune compromise or chronic intestinal diseases/conditions.

### Clinical data

A medical history was obtained from all study participants. CDI patients received phone follow ups every 14 days for 45 days, to assess for recurrent CDI (initial cessation of diarrhea, followed by recurrence within 45 days from completion of therapy and a positive stool test for *C. difficile*). For disease severity classification, individuals with new onset CDI were given an ATLAS score using published criteria ^15^. Individuals with recurrent CDI were classified as mild if they never required hospitalization. If they required hospitalization or administration of IV fluids during a CDI episode, they were defined as moderate, and if they had multiple recurrent hospitalizations they were classified as severe.

### Sample collection and processing

Peripheral blood was collected into 9mL sodium heparin vacutainers (Becton-Dickinson (BD)) and transported at ambient temperature. OX40 assays were set up as described below using whole blood within 4h of collection and plasma aliquots were stored at -80°C.

### Reagents

Staphylococcal enterotoxin B (SEB; Sigma-Aldrich, St. Louis, USA) was used at 1μg/mL. Pediacel®, a pentavalent vaccine containing components of pertussis vaccine, diphtheria and tetanus toxoids, inactivated poliomyelitis vaccine and *Haemophilus influenzae* B conjugate vaccine (Sanofi Pasteur Ltd, Lyon, France) was used at 1/40 dilution. *C. difficile* TcdA and TcdB toxoids (formaldehyde inactivated; List Biological Laboratories Inc. Campbell, USA) were used at 10μg/mL and 5μg/mL respectively. Soluble anti-CD3 (clone OKT3, UBC Antibody Lab, Vancouver, Canada) was used at 0.25μμg/mL. Tetanus toxoid was used at 2μg/mL (Enzo Life Sciences, Inc, Farmingdale, NY, USA).

### C. difficile growth and TcdBCROPS protein production

*C. difficile* ATCC strain 9689 was grown on YT agar plates anaerobically for 48-72 hours at 37ºC then colonies were grown in BHI broth at 37°C in an anaerobic chamber for 24-48 hours. Genomic DNA was extracted as previously described ^16^ and TcdB^CROPS^ amplified using the following primers:

5’-GGTTCGAACTATTCACTAATCACTAATTGAGC - 3’ and

5’-CTCGGATCCTGAAGAAAATAAGGTGTCACAAG-3’.

TcdB^CROPS^ was expressed with a 6xHis tag in *Escherichia coli* BL21(DE3)PLysS and purified by metal affinity chromatography as previously described ^17^.

### Identification of antigen-specific T cells using the OX40 assay

The OX40 assay was performed as previously described ^14^. OX40 assay mAb panel is listed in **Supplemental Table 1**. A four-laser LSRII FortessaX20 flow cytometer (BD) was used for data acquisition, utilising application settings. Assay cut offs were >0.02% of CD4^+^ T cells (being the mean ± 3SD of unstimulated wells) and consisting of at least 20 cells. Analysis was performed using FlowJo software (v10.6.1 Treestar Inc, Ashland, USA). IBD cohort data analysis used the DownSample v3.0.0 plugin to generate populations of 100 cells from CD25^+^OX40^+^ populations, which were then concatenated and analysed using tSNE v2.0 ^18^ and FlowSOM v1.5 ^19^ plugins.

### Cytokine analysis

Cell supernatants were collected following either cell growth of 5h stimulation with 10ng/mL phorbol 12-myristate 13-acetate (PMA) and 500ng/mL ionomycin in presence of 10μg/mL Brefeldin A (Signa-Aldrich). Concentrations of secreted cytokines in cell supernatants were measured using the 13-plex Th cytokine bead array kit according to manufacturer’s directions (BioLegend). CBA data were acquired on a 3-laser Cytoflex cytometer (Beckman Coulter) and analyzed using FCAP array v3 software (BD).

### ELISA

Quantification of anti-TcdA, TcdB IgG and IgA levels in plasma was performed by ELISA as previously described ^14^, using plates coated with 1μg/mL of TcdA or TcdB toxoids from List Biological Laboratories Inc (Campbell, CA, USA) or 1/100 dilution of Pediacel. Levels of anti-TcdB^CROPS^ IgG and IgA were measured by ELISA using 1μg/mL. A standard made from pooled plasma samples from subjects in our cohorts with high-titre antibodies was included with each ELISA and used to generate relative arbitrary units (AU) for cross-assay standardization.

### Isolation of TcdB-specific CD4^+^ T cells

Approximately 30mL of peripheral blood was centrifuged at 500 x g for 15min with brake off and 5mL of the buffy coat layer collected. This was mixed with 3mL Iscove’s Modified Dulbecco’s Medium (Thermo Fisher Scientific, Waltham, USA) and 4mL plated per well in a 6-well tissue culture plate (BD) and stimulated with 5μg/mL TcdB for 40-48h. CD4^+^ T cells were isolated using the CD4^+^ RosetteSep enrichment cocktail (STEMCELL Technologies) and TcdB-specific live CD4^+^CD25^+^OX40^+^ were isolated using a four-laser FACSAria IIu cell sorter (BD) to >95% purity.

### T cell expansion

Sorted cells were expanded with autologous APCs and antigen (or soluble anti-CD3) in presence of 200U/mL IL2 (Proleukin). APCs were obtained from PBMCs by negative isolation with CD3^+^ EasySep kit (Miltenyi Biotec) and irradiated 25Gy. Irradiated APCs were plated at 1 x 10^5^ cells/well in U-bottom 96 well plates and 20,000 sorted CD4^+^ T cells added. Antigens were added at specified concentrations and cells incubated at 37°C (5% CO^2^). T cells were restimulated every 14 days. Culture media for all T cell culture experiments was X-VIVO 15 (Lonza) supplemented with 5% heat-inactivated human serum (NorthBio Inc, Toronto, Canada), 1% Glutamax and 1% Penicillin-Streptomycin (Invitrogen). Cell proliferation was measured by staining with 5μM Cell Proliferation Dye eF450 (Invitrogen).

### HLA typing and TCR sequencing

HLA typing of the HLA-DRB1 locus was performed to a 4-digit resolution using sequence-specific oligonucleotide (SSO) typing by the Immunology Laboratory at Vancouver General Hospital. For TCR sequencing, DNA was extracted from isolated TcdB-specific CD4^+^ cells using QIAmp DNA mini kit according to manufacturers’ instructions (Qiagen). TCR sequencing of the beta chain was performed by Adaptive Biotechnologies (Seattle, USA) and analyzed using their online immunoSEQ analyzer software.

### Tetramer guided epitope mapping

Epitope mapping was performed as previously described 20 to identify HLA-DRB-restricted peptide epitopes within the CROPS domain of *C. difficile* toxin B (UniProt ID: P18177, amino acid positions 1831 – 2366) using a peptide library of 20mers overlapping by 12 amino acids (total of 66 peptides, Mimotopes, Australia). MP63, an influenza (flu) epitope, was used as a positive control tetramer ^21^. Briefly, PBMCs were stimulated with pools of 5 consecutive and overlapping peptides per well. After 14 days cells were stained with PE-conjugated patient-specific HLA-DRB1 tetramers loaded with a mix of the peptides from each pool. Cultures showing positive tetramer staining were further stained and analyzed with tetramers loaded with individual peptides to reveal the single peptide(s) recognized within the pool.

### Statistics

Statistical analyses between 2 groups used Mann-Whitney *U* test or, for paired samples, Wilcoxon signed rank test. Analysis of ≥ 3 groups used Kruskal-Wallis one-way ANOVA or, for paired samples, a Friedman one-way ANOVA with Dunn’s multiple comparison post-test. Correlation analyses calculated Spearman’s rho (r). P values were considered significant when < 0.05. Prism v8 (GraphPad Software Inc) was used for all statistical analyses. Error bars represent median ± interquartile range; *p ≤ 0.05, **p ≤ 0.01, ***p ≤ 0.001, ***p ≤ 0.0001 and ns = not significant.

## RESULTS

### Levels of C. difficile toxin-specific IgG do not distinguish infection from colonization

Studies of immunity to *C. difficile* have mostly focused on quantifying anti-toxin antibodies ^22^. Therefore, we asked whether plasma levels of anti-toxin antibodies could discriminate between healthy controls and recurrent or new onset CDI patients. ELISAs were used to measure TcdA and TcdB-specific IgG and IgA. We also measured antibodies specific for the C-terminal combined repetitive oligopeptides (CROPs) domain of TcdB (TcdB^CROPS^). This region is required for receptor binding and is the target of neutralizing antibodies ^23^. In all cohorts, levels of anti-TcdA IgG and IgA antibodies were significantly higher than anti-TcdB or anti-TcdB^CROPS^ (**Supplemental Figure 1**). When comparing between cohorts, only anti-TcdB IgA levels were significantly enriched in CDI patients (both recurring and new onset) compared to healthy controls (**Figure 1A)**.

**Figure 1.**
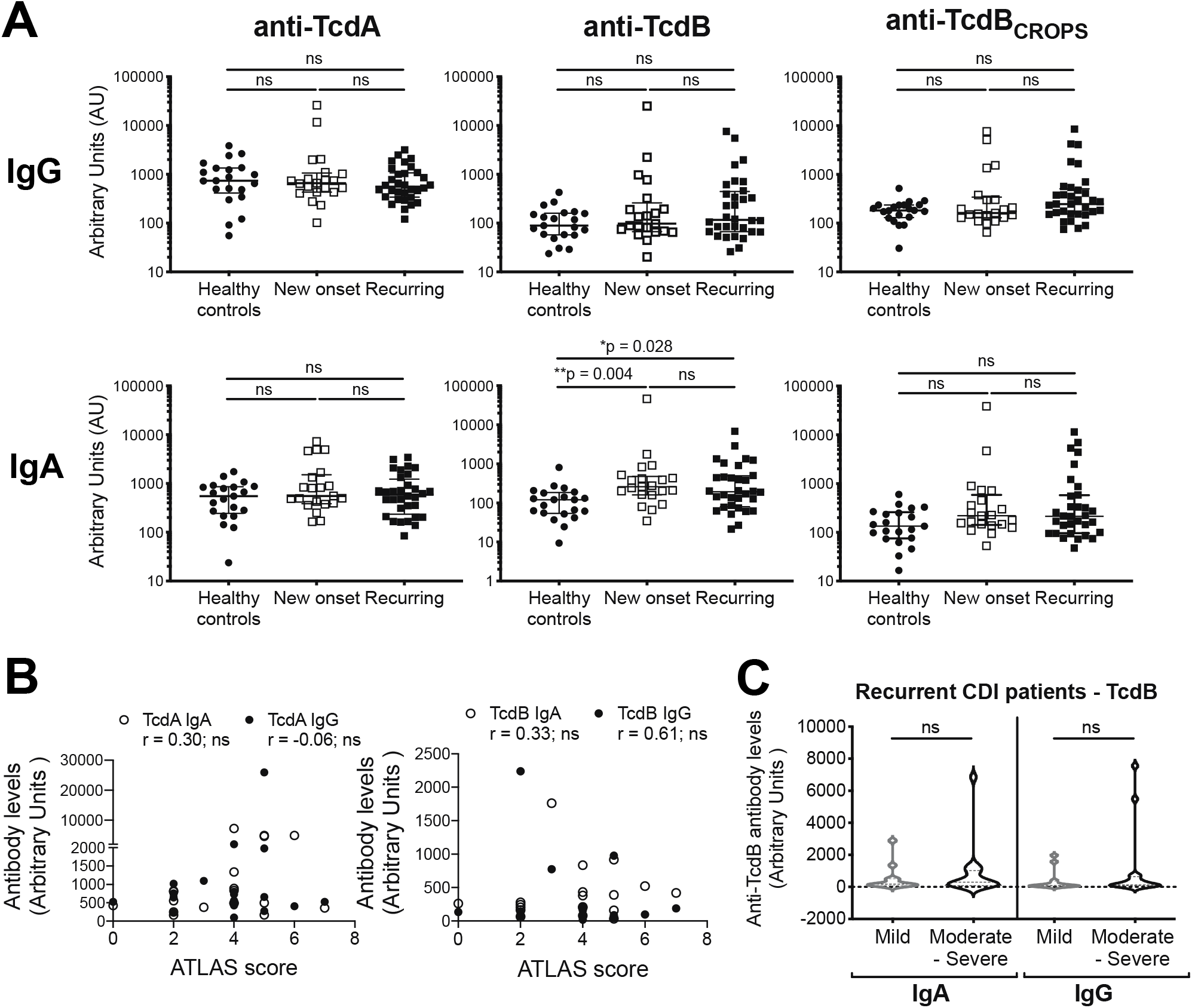
Anti-TcdB IgA responses but not IgG are elevated in CDI patients. **(A)** ELISAs were performed with plasma to assess levels of anti-TcdA, TcdB and TcdB^CROPS^ IgG and IgA for n=21 controls, n=32 recurring and n=21 new onset patients. Results are shown comparing levels of IgG (top row) and IgA (bottom row) between patient groups, using pooled high-titre plasma as internal control to obtain arbitrary units for comparison across samples. Kruskal-Wallis tests were performed. **(B)** Anti-TcdB and TcdA IgA and IgG levels did not correlate with ATLAS score in new onset patients (n=19; Spearman’s correlation). **(C)** Anti-TcdB IgG and IgA levels in recurrent patients with a history of mild (n=12) or moderate-severe illness (n=16; Mann-Whitney U tests).

We next asked if there was an association between antibody levels and disease severity. In the new onset cohort we used the ATLAS (Age, Treatment, Leukocyte count, serum Albumin, Serum creatinine) score as a measure of disease severity ^15^, and for the recurrent CDI cohort disease severity was determined using disease course (see methods). We found no associations with disease severity and antibody levels in either cohort (**Figure 1B-C**).

### Patients with active C. difficile infection have circulating toxin-specific CD4^+^ T cells

Having found that the magnitude of anti-toxin antibodies were not good indicators of disease, we utilized an antigen-induced marker assay, the ‘OX40 assay’ to quantify antigen-specific CD4^+^ T cells. This assay does not require knowledge of epitopes or HLA-restriction ^24–26^. Unfractionated peripheral blood was incubated with TcdA or TcdB toxoids from *C. difficile* ATTC strain 43255 for 44h and antigen-specific CD4^+^ T cells identified by induced co-expression of CD25 and OX40. Stimulation with Pediacel^?^, a pentavalent vaccine used in childhood vaccination, served as a positive control for antigen-specific T cell activation. Representative data from a subject with recurrent CDI are shown in **Figure 2A**, showing that induced-expression of CD25^+^OX40^+^ CD4^+^ T cells is detectable upon TcdA or TcdB stimulation.

**Figure 2.**
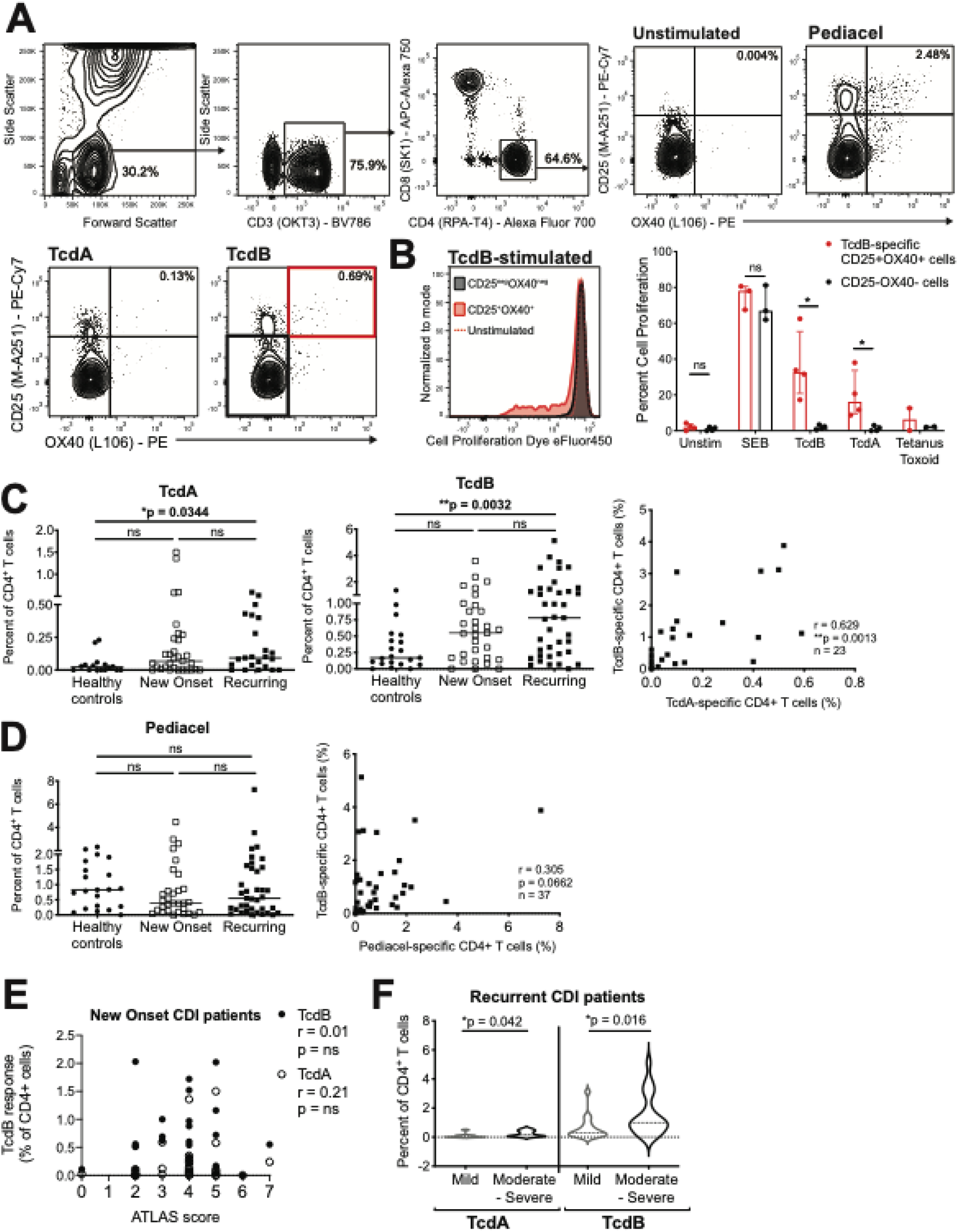
Proportions of anti-TcdA and TcdB CD4^+^ T cells increase with *C. difficile* infection. **(A)** Gating strategy for whole blood OX40 assays, and representative responses for unstimulated wells and wells stimulated with Pediacel, TcdA and TcdB. **(B)** *In vitro* autologous proliferation assays with sorted TcdB-specific CD4^+^CD25^+^OX40^+^ and non-TcdB-specific CD4^+^CD25^−^OX40^−^ T cells, with cells either left unstimulated (n=4) or stimulated with SEB (n=3), TcdB (n=4), TcdA (n=4) or tetanus toxoid (n=2). A representative proliferation plot is shown for a TcdB-stimulated assay; Mann-Whitney U tests performed. **(C)** CD4^+^ T cell specific for TcdA (controls n=16; new onset n=31; recurrent n=23) or TcdB (controls n=21; new onset n=31; recurrent n=39), Kruskal-Wallis tests; correlation analysis calculated Spearman’s rho (r) between these responses in recurrent patients (n=23). **(D)** Pediacel-specific CD4^+^ T cells (controls n=21; new onset n=31; recurrent n=37), Kruskal-Wallis tests; correlation analysis calculated Spearman’s rho (r) between TcdB and Pediacel responses in recurrent patients (n=37). **(E)** Proportions of TcdB and TcdA-specific CD4+ T cells correlated with ATLAS score in new onset patients (n=28); Spearman’s correlation; and **(F)** Proportions of TcdA and TcdB specific CD4+ T cells in recurrent patients with mild (n=18) or moderate-severe symptoms (n=21; Mann-Whitney U tests).

To confirm antigen specificity, CD25^+^OX40^+^ and CD25^−^OX40^−^ CD4^+^ T cells were sorted from TcdB-stimulated samples and re-stimulated with the super-antigen staphylococcus enterotoxin B (SEB; positive control), TcdB, TcdA, or tetanus toxoid from *Clostridium tetani*. Both cell populations proliferated in response to non-specific stimulation with SEB, but only the CD25^+^OX40^+^ cells proliferated in response to TcdB with some cross-reactivity to TcdA, and a small amount of proliferation to tetanus toxoid (**Figure 2B**).

To study the CD4^+^ T cell response to *C. difficile* toxoids in health versus CDI, we examined responses in our three cohorts (healthy controls, new onset CDI, and recurrent CDI). We found that the majority of healthy controls had detectable TcdB, but not TcdA, specific CD4^+^ T cells, and the proportion of these cells increased in both new onset and recurrent CDI. In 18/30 recurrent CDI patients, >1% of circulating CD4^+^ T cells were TcdB-specific, and there was a strong positive correlation between the presence of TcdB and TcdA-specific cells (**Figure 2C**). In contrast, there were no differences in the frequency of Pediacel-specific CD4^+^ T cells between the groups and no correlation between the presence of TcdB and Pediacel-specific CD4^+^ T cells (**Figure 2D**). These results indicate the increase in TcdA and TcdB-reactive CD4^+^ T cells is antigen and disease-specific.

We next asked whether the frequency of toxin-specific CD4^+^ T cells correlated with disease severity. We found no correlation between the ATLAS score and frequencies of TcdA or TcdB-specific CD4^+^ T cells in the new onset cohort (**Figure 2E**). For the recurrent CDI cohort, the frequency of TcdA, and to a greater extent, TcdB-specific CD4^+^ T cells was significantly higher in patients with moderate-severe disease, compared to those with mild disease **(Figure 2F**). Although TcdB-specific CD4^+^ T cells are present in most individuals, their frequency is significantly increased with repeated and more severe *C. difficile* infection. Thus, toxin-specific CD4^+^ T cells, but not antibodies, may be a useful biomarker of disease severity.

### Subjects with recurring CDI have a reduced frequency of TcdB-specific CD4^+^ Th17 cells

Having shown that TcdB-specific CD4^+^ T cells are readily detected in peripheral blood, we next examined their phenotype in more detail to quantify proportions of Th1, Th2, Th17 and Th17.1 cells on the basis of characteristic chemokine receptor expression (**Figure 3A**). We found that TcdB-specific CD4^+^ T cells were enriched for Th17 cells and that the proportion of Th17 cells was specifically reduced in patients with recurring CDI (**Figure 3B**). This reduction in Th17 cells in recurrent CDI subjects was unique to TcdB as no differences in T cell phenotype/proportions were observed in response to the vaccine antigen Pediacel (**Figure 3B**).

**Figure 3.**
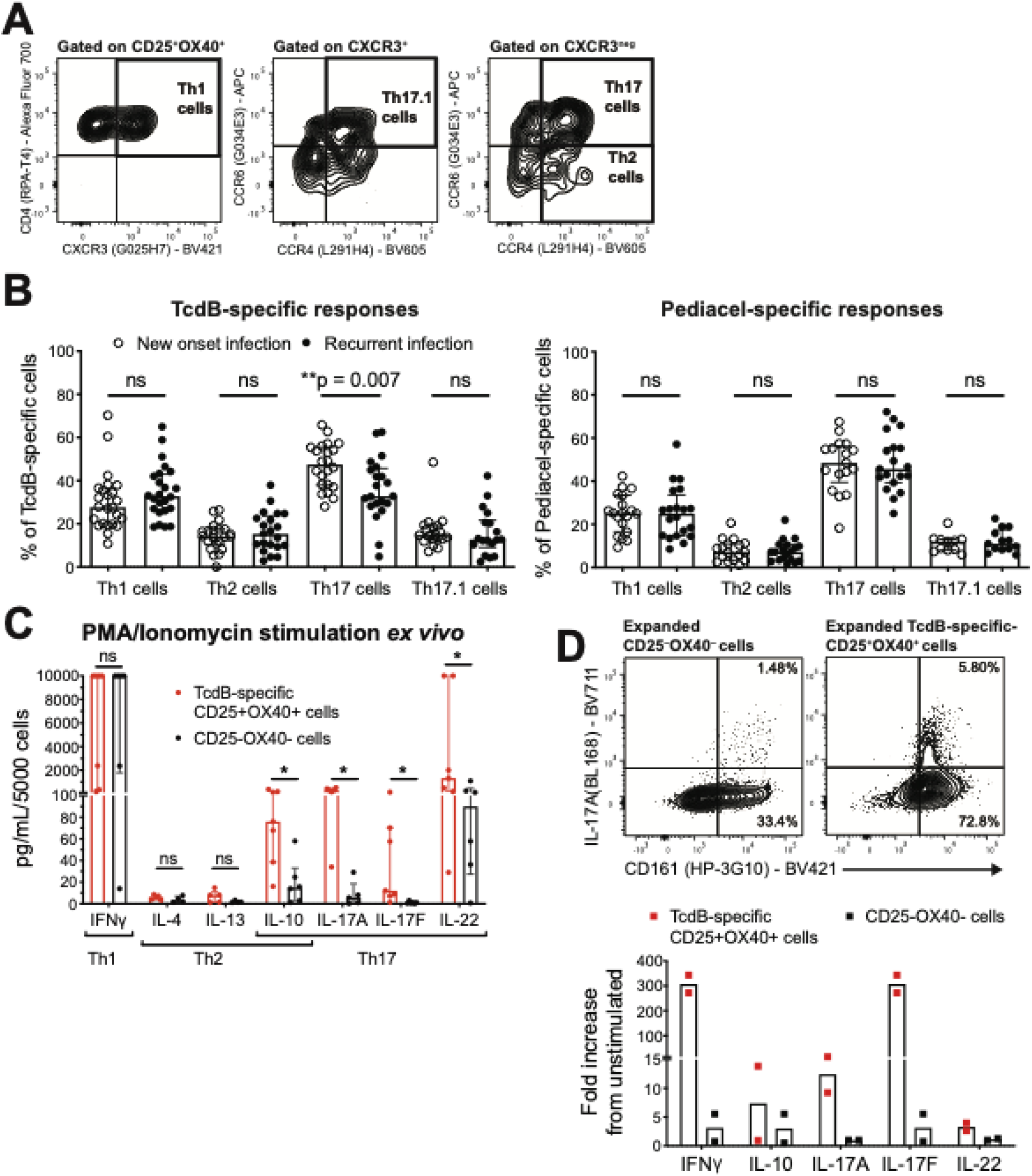
TcdB-specific CD4^+^ T cells are enriched for Th17 cells, which are reduced in recurrent infection. **(A)** Gating strategy for phenotypic analysis of TcdB-specific CD4^+^CD25^+^OX40^+^ T cells. **(B)** Comparison of proportions of Th1, Th2, Th17 and Th17.1 cells within TcdB or Pediacel-specific CD4^+^ T cells in recurrent versus new onset CDI patients. Kruskal-Wallis tests were performed. **(C)** Cytokine secretion of sorted TcdB-specific CD4^+^CD25^+^OX40^+^ and non-TcdB-specific CD4^+^CD25^−^OX40^−^ T cells following PMA/Ionomycin stimulation (n=6 recurrent CDI patients). **(D)** Sorted cells from (C) were expanded with autologous presentation of TcdB for 5d; representative staining (from n=3) shown of CD161 expression and IL17A secretion following addition of brefeldin A. Amounts of cytokine in supernatant were measured from wells without brefeldin A added, n=2.

To confirm the functional bias towards Th17 cells, CD25^+^OX40^+^ and CD25^−^OX40^−^ CD4^+^ T cells were sorted from TcdB-stimulated samples, re-stimulated with PMA and ionomycin, and secreted cytokines analysed. Consistent with the cell surface phenotype data, TcdB-specific CD25^+^OX40^+^ cells secreted significantly higher levels of the Th17 cell-associated cytokines IL10, IL17A, IL17F and IL22 compared to CD25^−^OX40^−^ CD4^+^ T cells (**Figure 3C**). In contrast, there were no differences in levels of the Th1 cell cytokine IFNγ, or the Th2 cytokines IL4 and IL13. The Th17-bias of TcdB-specific CD4^+^ T cells was further confirmed by intracellular cytokine staining following *in vitro* expansion showing these cells expressed more of the Th17 cell marker CD161 and secreted more IFNγ IL10. IL17A, IL17F and IL22 compared to CD25^−^OX40^−^ CD4^+^ T cells (**Figure 3D**). Thus, TcdB-specific CD4^+^ T cells are enriched for Th17 cells and their reduced proportions in patients with recurrent CDI suggests they are important for protective immunity. TcdB-specific Th17 cells also appear to be enriched for IL17A^+^IFNγ^+^ cells, a subset previously shown to be induced by fungal and bacterial pathogens ^27,28^.

### IBD patients with no history of CDI have TcdB-specific CD4^+^ T cells with an altered phenotype

Individuals with inflammatory bowel disease are at increased risk for CDI and experience more severe and recurrent disease ^29^, so we examined whether they may have underlying alterations in TcdB-specific CD4^+^ T cells. Blood was collected from a cohort of age- and sex-matched healthy controls, Crohn’s disease (CD) and ulcerative colitis (UC) patients. Clinical data for this cohort have been previously published ^14^; one UC subject with a recorded positive laboratory PCR test for *C. difficile* was excluded. We previously reported that T cell responses to Pediacel in both CD and UC patients were significantly reduced compared to healthy controls ^14^; therefore the frequency of TcdB-specific CD4^+^ T cells was normalized to account for the overall reduction in T cell memory responses in IBD patients. Upon normalization, we found the proportions of TcdB-specific CD4^+^ T cells were significantly higher in CD and UC patients compared to healthy controls (**Figure 4A**). We also observed a slight negative correlation with Harvey-Bradshaw index and the proportion of Integrin β7^+^ TcdB-specific CD4^+^ T cells, indicating that milder symptoms were associated with a higher proportion of gut homing TcdB-specific T cells in peripheral blood (**Figure 4B**).

**Figure 4.**
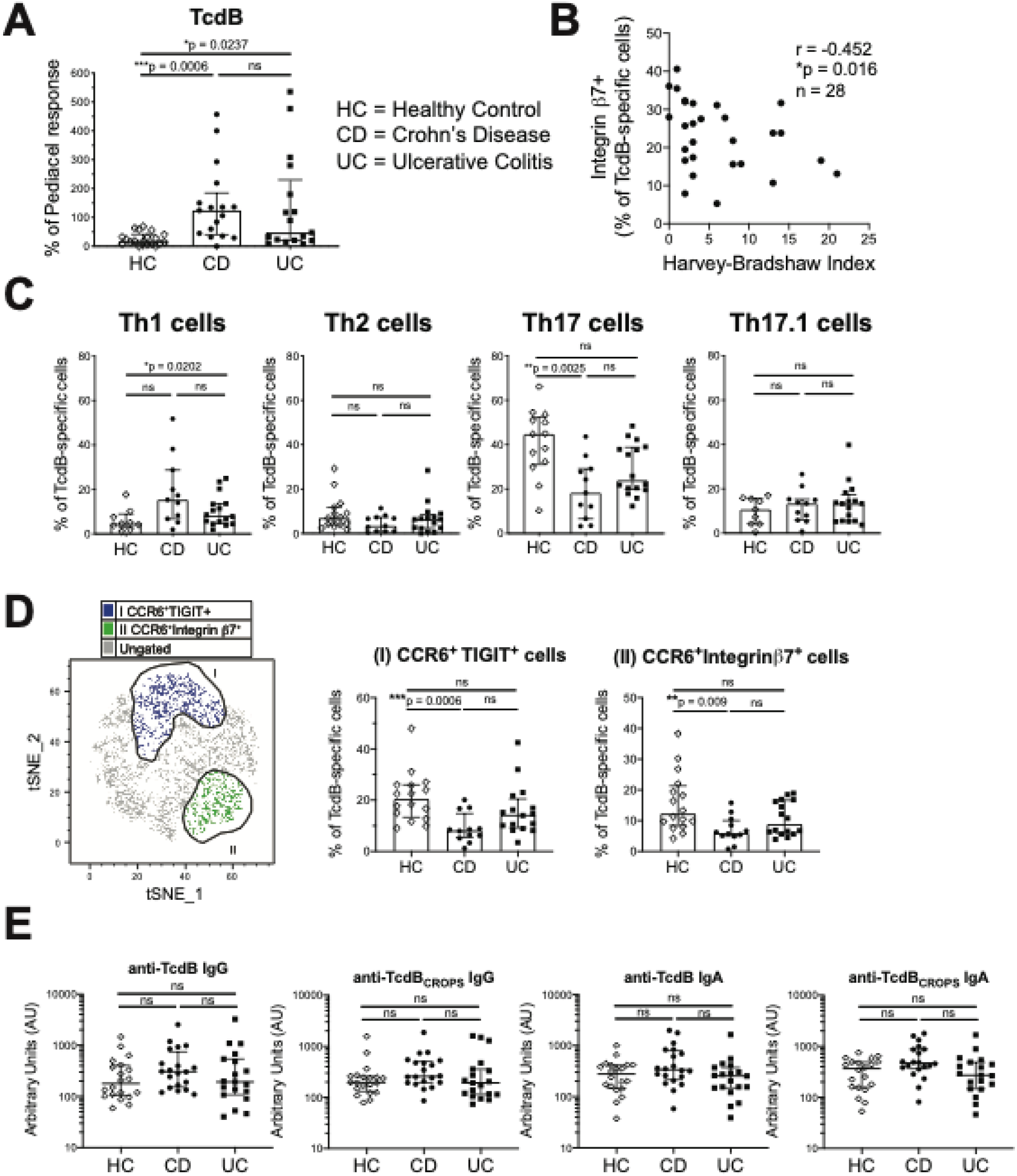
IBD patients have diminished proportions of Th17 TcdB-specific CD4^+^ T cells. TcdB and pediacel-specific CD4^+^ T cells were quantified in Crohn’s disease (CD, n=20), ulcerative colitis (UC, n=19) and healthy controls (HC, n=20). **(A)** TcdB responses were normalised as a percent of the Pediacel response. **(B)** Spearman’s rho correlation analysis between Harvey-Bradshaw index and the proportion of integrin β7^+^ TcdB-specific CD4^+^ T cells (n=28). **(C)** The proportion of Th1, Th2, Th17 and Th17.1 cells within TcdB-specific CD4^+^ T cells was measured using flow cytometry and the gating strategy in Figure 3. **(D)** Data from C were analyzed using unbiased clustering analysis using FlowSOM. Two clusters of TcdB-specific CD4+ T cells were found that discriminated between patient groups; clusters are shown overlaid on a tSNE plot. Frequencies of each identified cluster were confirmed by hierarchical gating within the TcdB-specific T cell gate, with proportions shown for all groups. Kruskal-Wallis tests were performed. **(E)** ELISA was used to measure the levels of anti-TcdB and -TcdB^CROPS^ IgG and IgA in all groups.

Similar to our findings in recurrent CDI patients, the proportion of TcdB-specific Th17 cells was decreased in both CD and UC patients compared to healthy controls (**Figure 4C**). There was also a significant increase in the proportions of Th1 cells in UC patients compared to healthy controls, but no differences in proportions of Th2 or Th17.1 cells. Dysfunctional TcdB-specific Th17 cell responses were further confirmed by FlowSOM unbiased clustering analysis. Within TcdB-specific cells, we found two clusters both expressing the Th17-associated marker CCR6 with proportional differences between IBD patients and healthy controls (**Figure 4D**). Cluster I expressed CCR6 and the co-inhibitory receptor TIGIT (T cell immunoreceptor with Ig and ITIM domains), and cluster II expressed CCR6 and integrin β7. Both these TcdB-specific Th17 cell sub-populations were significantly decreased in CD patients compared to healthy controls (**Figure 4D**).

Interestingly, there was no difference in the levels of anti-TcdB or anti-TcdB^CROPS^ IgG or IgA between IBD patients and controls (**Figure 4E**), further confirming the importance of measuring the frequency and phenotype of TcdB-specific CD4^+^ T cells rather than antibodies to measure immunity to *C. difficile*.

### The TcdB-specific TCR repertoire is polyclonal and includes HLA-DR7-restricted epitopes

A diverse TCR repertoire is important for effective immunity ^30^, but chronic antigen exposure, for example through reinfections, can narrow the repertoire ^31^. We investigated whether there was evidence of TCR repertoire bias, clonal expansion and/or public clonotypes within TcdB-specific CD4^+^ T cells by TCR variable β chain repertoire sequencing of sorted TcdB-specific CD4^+^ T cells for n = 5 recurrent CDI patients. All clonotype analysis used amino acid sequences. We found that TcdB-specific CD4^+^ T cells were polyclonal: > 75% of all productively rearranged templates had a unique clonotype and the Simpsons’ clonality index was < 0.03, indicating a low frequency of identical clonotypes (**Figure 5A**). The polyclonality of these responses was further illustrated by analysis of the Jaccard similarity index (100% overlap would be an index of 1, **Figure 5B**). We found several clonotypes were present in at least two individuals, possibly representing public clonotypes (**Figure 5B**). There was no bias in TCR beta chain variable region (TRBV) usage (**Figure 5C**).

**Figure 5.**
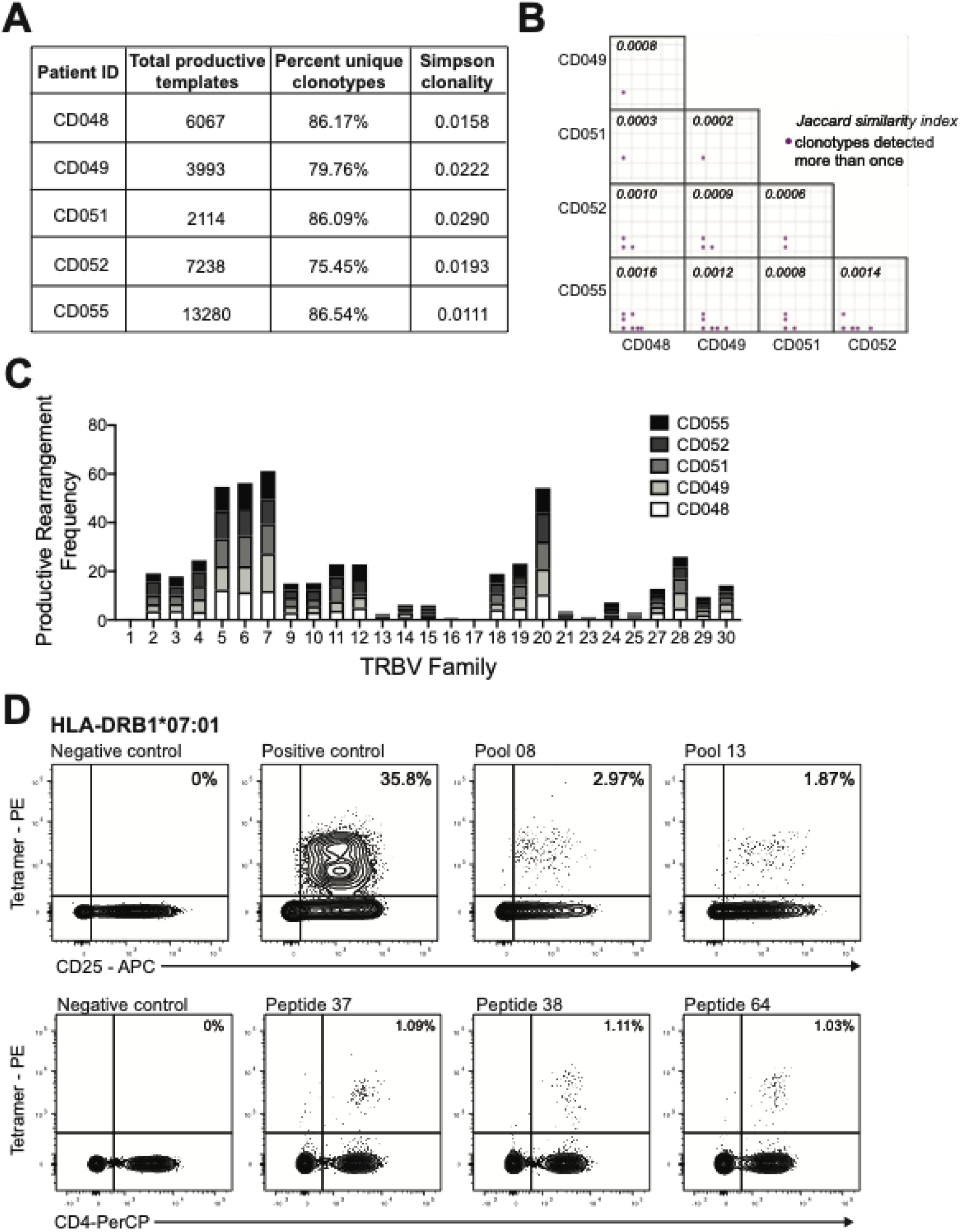
TcdB-specific CD4^+^ T cells are polyclonal TCR and include specificity for epitopes from TcdB^CROPS^. TcdB-specific CD4^+^ T cells were sorted from OX40 assays for n=5 recurrent CDI patients. The TCR repertoire was assessed by sequencing the beta chain. **(A)** The total productive templates sequenced, the percent that were unique clonotypes and the resulting Simpson’s clonality index are shown. **(B)** The similarity of samples was assessed using the Jaccard similarity index, with each dot representing a unique TCR clonotype. **(C)** The usage frequency of TRBV family gene segments is shown. **(D)** Tetramer-guided epitope mapping was used to identify HLA-DRB-restricted epitopes using peptide pools from the TcdB^CROPS^ region. Responses to the HLA-DRB1*07:01-restricted peptides 37, 38 and 64 were detected in n=2 recurring CDI patients.

To better define T cell epitopes, a peptide pool of 15mers overlapping by 12 amino acids from the TcdB ^CROPS^ domain was used for tetramer-guided mapping in HLA-DRB1*07:01-positive individuals. CD4^+^ T cells specific for three peptides (5’-3’): peptide 37 ‘VGFVTINDKVFYFSDSGIIE’ (aa2119 – 2138); peptide 38 ‘KVFYFSDSGIIESGVQNIDD’ (aa2127 – 2146); and peptide 64 ‘FTDEYIAATGSVIIDGEEYY’ (aa2335-2354) were detected in two subjects (**Figure 5D**). These epitopes may represent key immunogenic regions of TcdB and their discovery enables tetramer-based analysis of TcdB-specific CD4^+^ T cells in future studies. Tetramer based analysis may be particularly useful to detect functionally impaired or exhausted T cells, since such cells might not upregulate activation markers as efficiently.

### Fecal microbiota transplant increases TcdB-specific Th17 cells and antibodies

Some recurring CDI patients require FMT to cure infection, but how this treatment affects immunity to *C. difficile* was unknown. Blood from subjects with recurrent CDI was collected immediately prior to, and 8-12 weeks post, FMT. Clinical data showed that FMT was curative for 20/22 patients, as defined by absence of CDI recurrence without vancomycin prophylaxis for 3 months after FMT. T cell and antibody responses in the two patients who did not respond to FMT did not look different to the remainder of the cohort. Although the overall frequency of TcdB-specific CD4^+^ T cells was not altered by FMT, there was a significant increase in the proportion of TcdB-specific Th17 cells, and a small parallel decrease in Th2 cells (**Figure 6A**). Interestingly, there was a significant reduction in the frequency of Pediacel-specific CD4^+^ T cells post FMT, but no changes in the Th cell subsets proportions of these cells (**Figure 6B**). To rule out a global inhibition of T cell activation, we analysed responses to the super-antigen SEB. We found no change in frequency or phenotype of CD4^+^ T cells stimulated by SEB, supporting that changes in T cell frequency/function post-FMT are unique to their cognate antigen (**Supplemental Figure 2**).

**Figure 6.**
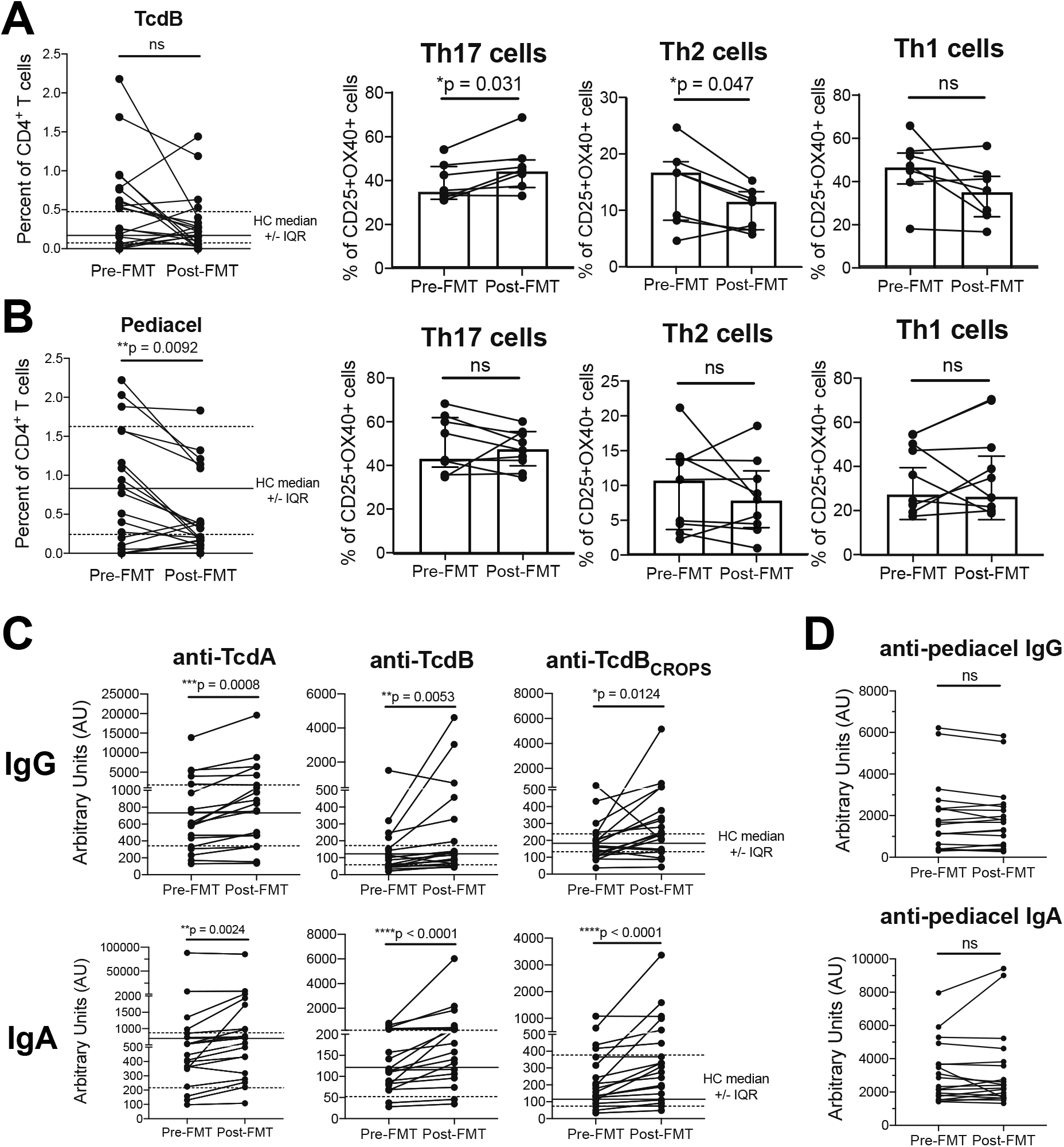
Proportions of Th17 TcdB-specific CD4^+^ T cell are increased post-FMT. Blood was collected from n=22 recurring CDI patients immediately prior to FMT and 8-12 weeks post-FMT. OX40 assays were performed and the magnitude and phenotype of responses are shown for **(A)** TcdB (phenotype data from n=7); and **(B)** Pediacel (phenotype data from for n=8). **(C)** Anti-TcdA, TcdB and TcdB^CROPS^; and **(D)** anti-Pediacel IgG and IgA levels were measured by ELISA for n=19 recurring CDI patients pre and post-FMT. Wilcoxon signed rank tests were performed. In (D) healthy control (HC) median (solid line) and interquartile range (dotted lines) are shown.

We found that anti-TcdA, TcdB and TcdB^CROPS^ IgA and IgG levels were all significantly increased post-FMT (**Figure 6C**). Importantly, anti-Pediacel IgG and IgA levels did not change post-FMT (**Figure 6D**), indicating that changes in pre-existing vaccine-generated immunity is not a global effect and may be restricted to circulating CD4^+^ T cells. Collectively, these results indicate that the positive effects of FMT may not only be related to changes in the microbiome, but also by improving CD4^+^ T cell and antibody mediated immunity to *C. difficile* toxins.

## DISCUSSION

There is an urgent need to understand immunity to *C. difficile* in order to advance treatment and diagnosis strategies for this common antibiotic-resistant bacterial pathogen. While most research has focused on humoral immunity, the CD4^+^ T cell response to *C. difficile* has remained unexplored, a critical missing piece due to the importance of CD4^+^ T cells in controlling antibody class-switching. We report here the first analyses of human CD4^+^ T cells with specificity to *C. difficile* toxins in healthy adults and patients with IBD or active CDI. We show that the T cell, but not antibody, response to TcdB distinguishes CDI and IBD patients from healthy controls and positively correlates with disease severity in recurrent CDI patients. Evidence that the significant reduction of TcdB-specific Th17 cells in recurrent CDI patients was improved following FMT therapy suggests that impaired TcdB-specific Th17 memory cells is an important aspect of CDI pathogenesis and may represent a novel therapeutic target.

Previous studies of CDI immunity have focused on antibodies and we confirmed reports showing IgA antibody responses to TcdA were higher than for TcdB ^32^. Anti-TcdB IgA, but not IgG, were significantly higher in both new onset and recurrent CDI patients compared to healthy controls, suggesting that infection results in increased production of IgA. For IgG, the lack of a difference between any of the cohorts suggests that these antibodies are generated following colonisation and are not increased during active infection. Evidence that IgG and IgA antibody responses to TcdB^CROPS^ were of similar magnitude to those against the entire TcdB protein indicates that the majority of anti-TcdB antibodies target the ^CROPS^ region, as previously suggested ^23^. The lack of association between IgG or IgA anti-toxin antibodies and disease severity in any patient group confirms previous reports that anti-toxin antibodies are not a useful diagnostic tool ^11,12^.

In contrast to the lack of differential antibody levels between patients and controls, we found that patients with either newly diagnosed or recurring CDI had higher frequencies of circulating CD4^+^ T cells specific for both TcdA and TcdB compared to healthy controls. Further, within recurrent CDI patients an increase in TcdA and TcdB-specific CD4^+^ T cells was associated with more severe disease. The finding of higher frequencies of TcdB-specific than TcdA-specific CD4^+^ T cells may correlate with the more prominent role of TcdB in disease pathogenesis ^33^. As there was no correlation between TcdB and Pediacel responses, the relatively high (i.e. ~1% of all circulating CD4^+^ T cell) frequency of TcdB-specific CD4^+^ T cells was likely not due to systemic immune activation, but rather reflective of a true expansion of these cells in active CDI.

We also investigated the Th cell composition of the TcdB-specific CD4^+^ T cells and found that, in CDI patients and healthy controls, these cells were enriched for cells with a Th17 cell phenotype. This finding echoes what has been shown for Th cell responses to other intestinal microbes, both pathogenic and commensal ^14,27,28,34,35^. Of note, the only other study to assess the phenotype of CD4^+^ T cells in CDI patients found increased IL17A^+^ cells in recurrent CDI compared to healthy controls, but did not examine antigen-specific responses ^36^. The importance of IL17 in the gut for protection against infections has been well-described ^34,35,37^, and our finding that TcdB-specific Th17 cells secrete both IL17A and IFNγ is reminiscent of *Staphylococcus aureus*-specific human Th17 cells, which are also enriched in dual-cytokine producing cells ^28^. Thus, reduced Th17 immunity appears restricted to TcdB-specific responses, although further comparison of these cells with healthy controls is required to assess functional differences.

Interestingly, we found that the proportion of TcdB-specific Th17 cells was significantly reduced in patients with recurring CDI compared to those with newly acquired CDI. In support of our data showing a beneficial role of Th17 cells in CDI, a recent study in mice found that IL17-producing γδ T cells protected against *C. difficile* infection ^38^. In humans, colonization with *C. difficile* within the first year of life is common but does not cause symptoms; however, after age two, CDI can occur in children with severe illnesses and/or heavy antibiotic use ^33^. Chen *et al* suggested that the increased proportions of IL17-producing γδ T cells in neonates could be a mechanism underlying their protection from CDI ^38^. A key area for future studies is the relationship between early life *C. difficile* colonisation, IL17-production, and the generation of Th17 memory to *C. difficile* antigens.

Although IBD patients are known to have increased risk of acquiring CDI, and of severe, complicated infections ^9^, routine screening for active CDI is not currently recommended, due to the inability to confidently distinguish colonization from disease ^10^. We found increased TcdB-specific CD4^+^ T cells in both Crohn’s disease and ulcerative colitis patients and, similar to our findings in CDI patients, TcdB-specific Th17 cells in Crohn’s disease patients were significantly reduced compared to healthy controls. These data support the idea that Th17 cells are beneficial for protective immunity to *C. difficile*. IBD patients also had reduced proportions of TcdB-specific Th17 cells that expressed integrin β7, a finding which correlated with more severe symptoms, suggesting that an ability to home to the gut is also important for protective immunity. Our data provide clues to an immunological mechanisms underlying the failure of drug interventions targeting IL17A in IBD patients ^39^ and indicate the overgrowth of *C. difficile* and low-level toxin production may be contributing to dysbiosis and gastrointestinal symptoms in these patients.

Our finding that a significant proportion of healthy controls with no history of CDI have CD4^+^ T cells specific to TcdB, and IgG and IgA specific to TcdA are consistent with published observations that environmental exposure to toxigenic *C. difficile* is common in the general population ^40^. TcdA and TcdB proteins are 63% homologous ^41^; therefore, our observation of TcdB-specific CD4^+^ T cells exhibiting cross reactivity to TcdA is not surprising and may indicate these responses are directed against shared homology regions. We identified that the TCR repertoire of TcdB-specific CD4^+^ T cells was polyclonal, similar to the Th17-biased response to *Candida albicans* ^27^. There were a few clonotypes that were observed in more than one individual, suggesting the presence of public TCR clonotypes within the anti-TcdB CD4^+^ T cell repertoire. We also performed class-II tetramer-guided epitope mapping of the TcdB ^CROPS^ domain and identified three DR7-restricted epitopes recognised in two recurrent CDI patients. Two of these TcdB^CROPS^ epitopes are overlapping peptides (peptides 37 and 38) that have consensus regions with the TcdA ^CROPS^ domain ^42^, potentially a region enriched for HLA-II-restricted immunodominant epitopes.

We had the opportunity to examine immune responses in patients before and after FMT for recurrent CDI. Surprisingly, we found that post-FMT patients had an increased proportion of TcdB-specific Th17 cells and anti-toxin antibodies. These findings are consistent with reduced TcdB-specific Th17 cells being associated with more severe disease outcomes. Interestingly, we also detected reduced CD4^+^ T cell responses to Pediacel post-FMT, although plasma antibody levels were unchanged from pre-FMT, indicating that FMT may trigger alterations in pre-existing memory CD4^+^ T cells. Whether the detected changes are due to alterations in the microbiota from FMT remains unknown. All patients were maintained on low-dose suppressive vancomycin prior to FMT as part of standard of care; therefore, it is possible that some of the immune changes were driven by microbiota restitution resulting from withdrawal of vancomycin. Other possible contributory factors include changes in diet or activity following FMT. It is interesting to note that 8 weeks following successful FMT, circulating levels of all measured anti-toxin antibodies increased. These results are potentially important for disease monitoring strategies and could form the basis of future studies to identify patients at risk of recurrent disease who may benefit from additional treatments or prophylaxis.

In conclusion, this is the first study investigating CD4^+^ T cell immunity generated following natural infection with *C. difficile*, revealing strong responses directed against the secreted toxins TcdA and TcdB, with a predominance of Th17 cells. The finding that FMT modulates the frequencies of these cells in parallel to curing disease suggests that strategies to prevent and/or treat CDI should investigate whether enhancing IL17/Th17 pathway activity could be a new therapeutic approach in this disease.

## Data Availability

There are no large datasets in this paper.

## ACKNOWLEDGMENTS

The authors wish to thank Mr. Matthew Suzuki at the Vancouver Gastrointestinal Research Institute for patient recruitment and sample collection; and Dr. Simon Hirota at University of Calgary for assistance with protocols for growing *C. difficile*.

## Author contributions

LC designed experiments, acquired, analysed and interpreted data and wrote the manuscript; WDR performed all ELISA experiments and data analysis and generated the TcdB^CROPS^ antigen, MQW designed experiments, acquired and analysed data; XW acquired and analyzed data; HP, LO, TL, RM and BB contributed to study design, patient recruitment, and critical revision of manuscript; RG, IC, EAJ and WWK performed tetramer-guided epitope mapping and critically revised the manuscript; MKL and TSS obtained funding and contributed to study concept, design, supervision, and critical revision of manuscript.

## Funding

Supported by a Canadian Institutes of Health Research Project Grant (20R76508 to M.K.L and T.S.S) and an Antimicrobial Resistance: Point of Care Diagnostics in Human Health grant (20R75988 to L.C, M.K.L and T.S.S); a grant from the Broad Medical Research Program at the Crohn’s & Colitis Foundation of America (IBD-0326 to M.K.L and T.S.S); a MISP grant from Merck Canada to T.S.S; a University of British Columbia Four Year Doctoral Fellowship (W.D.R), and the BC Children’s Hospital Research Institute Bertram Hoffmeister Postdoctoral Fellowship (L.C).

**Abbreviations**
CDI: *C. difficile* infection
FMT: Fecal microbiota transplant
IBD: Inflammatory bowel disease
CD: Crohn’s disease
UC: Ulcerative colitis
TCR: T cell receptor

## SUPPLEMENTAL DATA

**Supplemental Figure 1.**
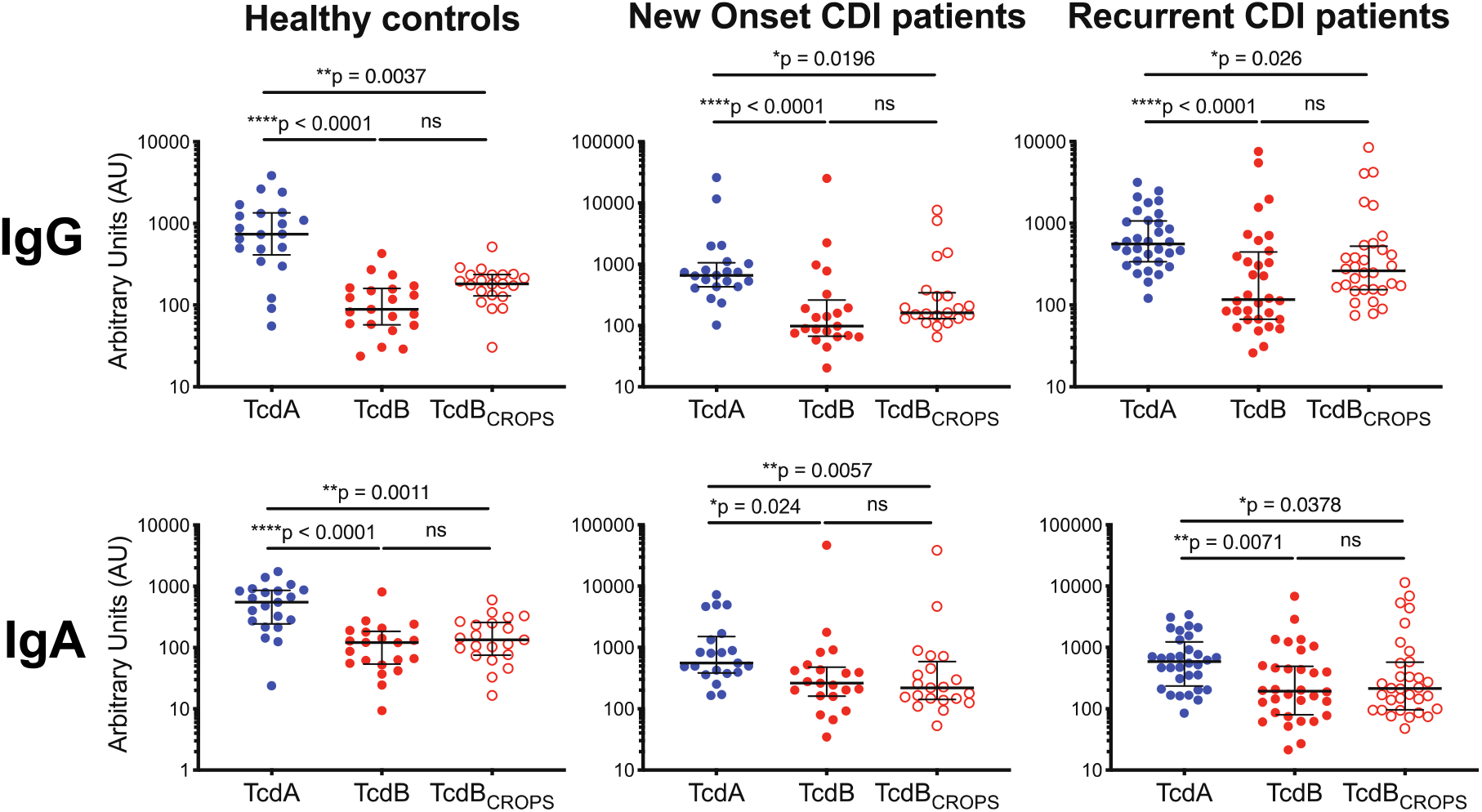
Comparison of anti-TcdA/TcdB IgG and IgA levels. ELISAs were performed with plasma to assess levels of anti-TcdA, TcdB and TcdB^CROPS^ IgG and IgA for n=21 controls, n=32 recurring and n=21 new onset patients. Results are shown comparing levels of anti IgG (top row) and IgA (bottom row) by target antigen for each patient group; Kruskal-Wallis tests were performed.

**Supplemental Figure 2.**
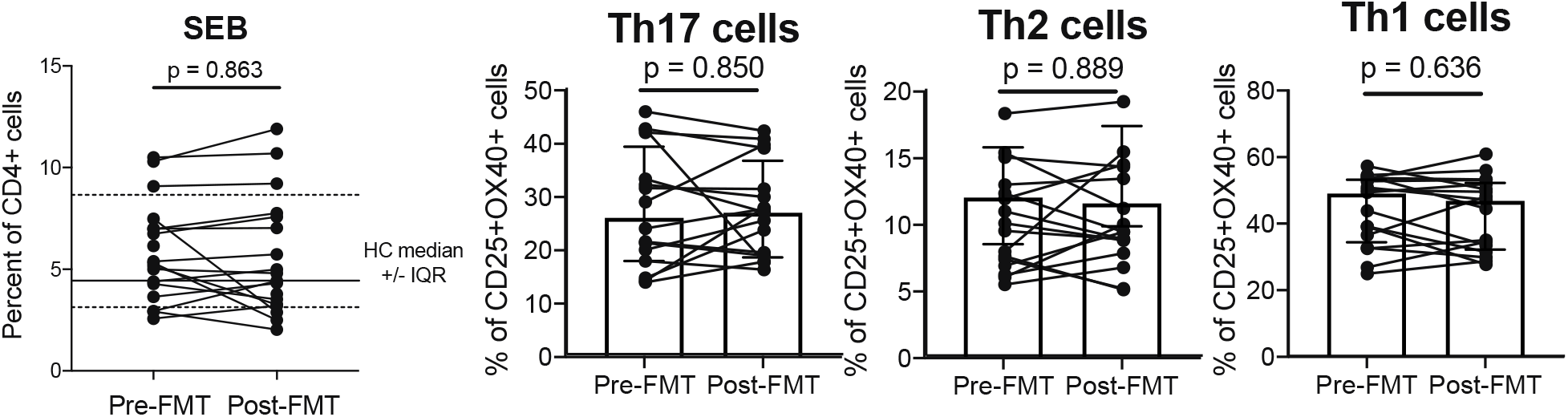
SEB-mediated T cell stimulation in recurrent CDI patients pre- and post-FMT. Blood was collected from n=22 recurring CDI patients immediately prior to FMT and 8-12 weeks post-FMT. OX40 assays were performed and the proportion and phenotype of SEB-stimulated T cells analyzed by flow cytometry (phenotype data from n=16).

**Supplemental Table 1.** mAb panel used for OX40 assay analysis.

| Specificity | Conjugation | Clone | Amount/test | Supplier | Cat number |
| --- | --- | --- | --- | --- | --- |
| CD3 | Brilliant Violet 785 | OKT3 | 0.5μL | BioLegend | 317330 |
| CD4 | Alexa Fluor 700 | RPA-T4 | 1μL | BD | 557922 |
| CD8 | APC-Fire750 | SK1 | 0.5μL | BioLegend | 344745 |
| CD25 | PE-Cy7 | M-A251 | 5μL | BD | 557741 |
| CD134(OX40) | PE | L106 | 20μL | BD | 340420 |
| CD39 | FITC | A1 | 2μL | BioLegend | 328206 |
| Integrin β7 | PE-Cy5 | FIB504 | 2μL | BD | 551059 |
| CXCR3 | Brilliant Violet 451 | G025H7 | 2μL | BioLegend | 353716 |
| CCR6 | APC | G034E3 | 2μL | BioLegend | 353415 |
| CCR4 | Brilliant Violet 605 | L291H4 | 1μL | BioLegend | 359418 |

